# Serological detection of SARS-CoV-2 IgG using commercially available immunoassays on dried blood spots collected from patients

**DOI:** 10.1101/2021.08.11.21261877

**Authors:** Gregory J Walker, Rebecca Davis, Zin Naing, Brad McEntee, Yonghui Lu, Tatijana Denadija, William D Rawlinson

## Abstract

Serological testing for SARS-CoV-2 antibodies provides important research and diagnostic information relating to COVID-19 immune response and surveillance. A major challenge when addressing protection post-infection or vaccination is the difficulty of specimen collection from infants and children. Dried blood spots (DBS) collected by finger prick or heel prick are a minimally invasive sample collection alternative previously used to detect antibodies to other viruses. In this study we evaluated DBS for the detection of SARS-CoV-2 antibodies on three commercially available enzyme (EIA) and chemiluminescent (CLIA) immunoassays by analysing paired DBS and serum samples collected from 54 subjects. We demonstrate that testing of DBS samples was highly sensitive and specific, and quantitative results strongly correlated with those of paired serum. These results suggest that DBS derived blood is a viable alternative to plasma or serum for use in EIAs and CLIAs, and has particular utility as a minimally invasive collection tool for COVID-19 serological testing of infants and children.

---

Serological methods for SARS-CoV-2 antibody detection are routinely performed on plasma or serum from peripherally collected venous blood. This can be inconvenient and often unacceptable to children. Dried blood spots (DBS) collected by finger prick or heel prick are a minimally-invasive alternative previously used to detect antibodies against Epstein-Barr, HIV, Hepatitis, Rubella and other viruses (1). Evaluation of DBS for the detection of SARS-CoV-2 antibodies on commercially available enzyme (EIA) and chemiluminescent (CLIA) immunoassays was performed by analysing paired DBS and serum samples from 54 subjects aged between 5 and 73 years.

Whole blood was collected from patients via venepuncture and spotted onto DBS cards (Whatman 903 Protein Saver), with the remainder put into a serum tube. DBS were then processed (Supplementary materials) and tested in parallel with matched sera for antibodies against SARS-CoV-2 using the spike-based ARCHITECT SARS-CoV-2 IgG II Quantitative Assay (Abbott Laboratories, Abbott Park, IL, USA), and the S1 spike- and nucleocapsid-based Euroimmun Anti-SARS-CoV-2 ELISA (Euroimmun, Lübeck, Germany) assays, following manufacturer’s instructions. The ARCHITECT SARS-CoV-2 IgG II Quantitative Assay records data as arbitrary units (AU), and these results were interpreted as positive (≥50 AU/mL) and negative (<50 AU/mL). For Euroimmun assays, the ratio of sample absorbance over the calibrator (OD ratio) was used to categorise sample results as positive (≥1.1), equivocal (1.1> OD ≥0.8), and negative (<0.8). Samples returning an equivocal result were excluded from sensitivity and specificity calculations in this analysis. Sensitivity and specificity were calculated using results of parallel testing on peripherally collected serum samples as the reference standard. Concordance between quantitative measurements were calculated with Pearson’s Correlation Coefficient.

For the spike-based ARCHITECT and Euroimmun assay there were no discordant results between any of the 54 paired samples. On both assays DBS testing displayed 100% sensitivity and 100% specificity with reference to matched serum. The correlation coefficients between quantitative serum and DBS measurements were r= 0.9662 (0.9421 – 0.9803, p<0.0001) and r= 0.9831 (0.9708 -0.9902, p< 0.0001), respectively (Figure). On a subset of 46 matched samples tested on the Euroimmun nucleocapsid-based assay there was one discordant result, and DBS testing exhibited 100% sensitivity. One of 5 pairs that had a negative result on serum testing was positive on DBS testing. The correlation coefficient between quantitative serum and DBS measurements were r= 0.9075 (0.8380 – 0.9480, p<0.0001).

**Figure.**
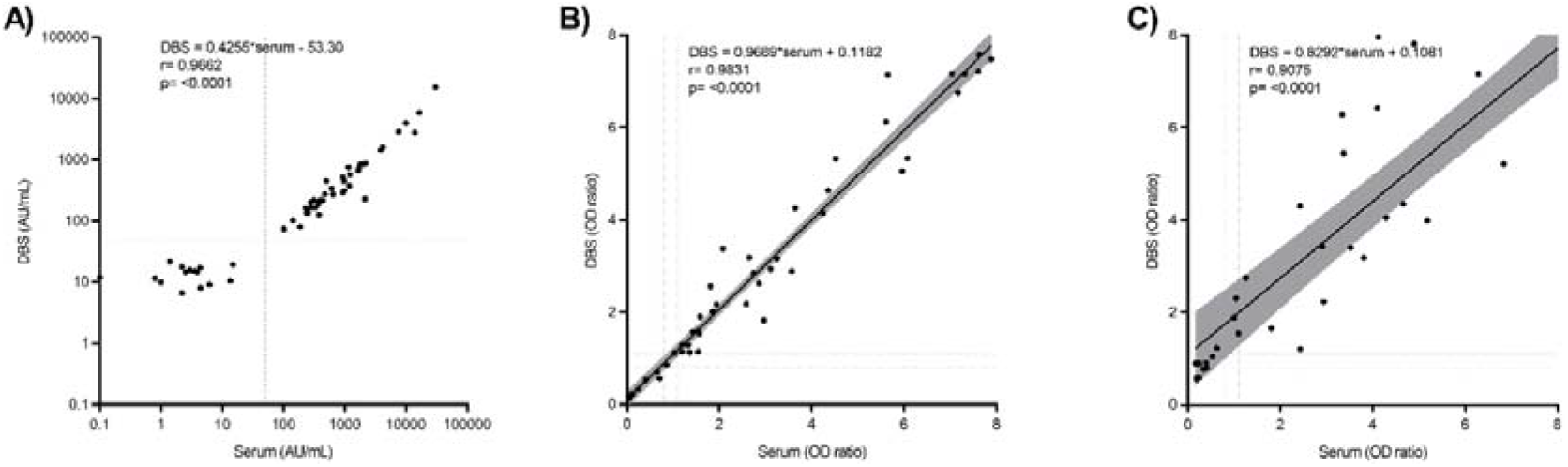
Linear regression of CLIA arbitrary units per millilitre (AU/mL) or EIA optical density (OD) ratios for the detection of anti-SARS-CoV-2 IgG in 54 paired dried blood spot (DBS) and serum samples. Samples were run on the ARCHITECT SARS-CoV-2 IgG II Quantitative Assay **(A)**, and the Euroimmun Anti-SARS-CoV-2 ELISA S1 spike **(B)** and nucleocapsid assays **(C)**. Dotted lines indicate the positive/negative, negative/equivocal, and equivocal/positive cutoffs of respective assays.

The high sensitivity and specificity of DBS tests and strong quantitative relationship with paired serum on the spike-based ARCHITECT CLIA and Euroimmun EIA demonstrates the validity of DBS collection for the detection of SARS-CoV-2 antibodies on these assays. This is the first study to validate the quantitative ARCHITECT assay for DBS testing, and the first to include paediatric specimens in analysis (Supplementary materials). Our findings also confirm those of recent studies in which adult DBS were validated using semi-quantitative EIAs targeting the S1 spike (2-4) and nucleocapsid proteins (3-5). These data suggest DBS derived blood is a viable alternative to plasma or serum collection. It has particular utility for testing infants and children, and in hotel or home quarantine where sample collection is performed away from clinical settings. It can reduce transmission risks to staff collecting samples from infected patients, as less time needs to be spent with the child to obtain the sample. Additionally, this non-invasive tool collection tool is well suited to COVID-19 surveillance.

## Data Availability

Data will be made available upon contacting the authors

## Supplementary Material

### Processing of dried blood spot samples

Whole blood was collected from patients via venepuncture and spotted onto Whatman 903 Protein Saver DBS cards. DBS samples were then processed by methods applicable to each assay platform.

For the ARCHITECT SARS-CoV-2 IgG II Quantitative Assay (Abbott Laboratories, Abbott Park, IL, USA) a 6mm spot was punched using a standard office hole punch and transferred to a 2 ml round bottom centrifuge tube. After 300µl of 0.25% Tergitol 15-S-9 in PBS was added, the sample tube was incubated on an orbital shaker at 400 rpm for 60 min, followed by a centrifugation at 10,000g for 10 min. Supernatant (130µL) was then transferred to a sample cup and loaded onto ARCHITECT i2000SR analyzer for testing. As the current on-market ARCHITECT SARS-CoV-2 IgG II Quantitative Assay is not claimed for DBS use, a Research Use Only (RUO) assay file was installed to facilitate the DBS sample measurement. The DBS method for this platform may be further modified to improve agreement with quantitative serum results.

For the Euroimmun Anti-SARS-CoV-2 ELISA (Euroimmun, Lübeck, Germany) assays, a single 4.76mm disk was punched and incubated in 250µL of ELISA buffer (kit component) for one hour at 37°C. The eluted solution was then pipette mixed thoroughly and 100uL was taken forward for analysis as per the manufacturer’s instructions.

**Table S1.** Age distribution of patients sampled

| Age | n= |
| --- | --- |
| All | 54 |
| 5-12 | 4 |
| 13-18 | 4 |
| 19-40 | 29 |
| 41-60 | 12 |
| >60 | 5 |

